# Feasibility of an Adaptive E-Learning Environment to Improve Provider Proficiency in Essential and Sick Newborn Care in Mwanza, Tanzania

**DOI:** 10.1101/2023.07.11.23292406

**Authors:** Peter Meaney, Adolfine Hokororo, Hanston Ndosi, Alex Dahlen, Theopista Jacob, Joseph R Mwanga, Florence S Kalabamu, Christine Joyce, Rishi Mediratta, Boris Rozenfeld, Marc Berg, Zack Smith, Neema Chami, Namala P Mkopi, Castory Mwanga, Enock Diocles, Ambrose Agweyu

## Abstract

**Introduction:** To improve healthcare provider knowledge of Tanzanian newborn care guidelines, we developed adaptive Essential and Sick Newborn Care (aESNC), an adaptive e-learning environment (AEE). The objectives of this study were to 1) assess implementation success with use of in-person support and nudging strategy and 2) describe baseline provider knowledge and metacognition.

**Methods:** 6-month observational study at 1 zonal hospital and 3 health centers in Mwanza, Tanzania. To assess implementation success, we used the RE-AIM framework and to describe baseline provider knowledge and metacognition we used Howell’s conscious-competence model. Additionally, we explored provider characteristics associated with initial learning completion or persistent activity.

**Results:** aESNC reached 85% (195/231) of providers: 75 medical, 53 nursing, and 21 clinical officers; 110 (56%) were at the zonal hospital and 85 (44%) at health centers. Median clinical experience was 4 years [IQR 1,9] and 45 (23%) had previous in-service training for both newborn essential and sick newborn care. Efficacy was 42% (SD±17%). Providers averaged 78% (SD±31%) completion of initial learning and 7%(SD±11%) of refresher assignments. 130 (67%) providers had ≥1 episode of inactivity >30 day, no episodes were due to lack of internet access. Baseline conscious-competence was 53% [IQR:38-63%], unconscious-incompetence 32% [IQR:23-42%], conscious-incompetence 7% [IQR:2-15%], and unconscious-competence 2% [IQR:0-3%]. Higher baseline conscious-competence (OR 31.6 [95%CI:5.8, 183.5) and being a nursing officer (aOR: 5.6 [95%CI:1.8, 18.1]), compared to medical officer) were associated with initial learning completion or persistent activity.

**Conclusion:** aESNC reach was high in a population of frontline providers across diverse levels of care in Tanzania. Use of in-person support and nudging increased reach, initial learning, and refresher assignment completion, but refresher assignment completion remains low. Providers were often unaware of knowledge gaps, and lower baseline knowledge may decrease initial learning completion or activity. Further study to identify barriers to adaptive e-learning normalization is needed.

**Key questions:** *What is already known on this topic*.

*Summarize the state of scientific knowledge on this subject before you did your study and why this study needed to be done*.

- In sub-Saharan Africa, gaps in care quality may contribute to its high neonatal mortality.
- Provider knowledge is a main driver of care quality, but current conventional in-service education methods are inadequate in adaptivity, reach, effectiveness, and refresher assignments.
- Hard copies of national guidelines have been disseminated to health facilities expectations are HCPs will learn and adhere to them.
- Adaptive eLearning, a subdomain of e-learning, holds the potential to overcome limitations to in-service medical education, but the optimal implementation strategy is unknown.

*What this study adds*.

*Summarize what we now know because of this study that we did not know before*.

- Baseline knowledge of essential and sick newborn care was low, mostly due to unconscious incompetence (providers thinking they were correct when they were incorrect).
- Initial learning completion increased significantly with the use of an in-person program manager and an escalating nudging strategy, and technical issues were not identified as a significant limitation to participation.

*How this study might affect research, practice, or policy*.

*Summarize the implications of this study*.

- Provider self-reporting may underestimate knowledge gaps as most gaps are not known by providers.
- Adaptive e-learning may be a feasible and acceptable way to disseminate guideline and improve quality of care if an implementation strategy can be identified to increase refresher assignment completion.
- Once the ideal implementation strategy is identified, effectiveness of adaptive e-learning at scale can be evaluated.

## INTRODUCTION

The Government of Tanzania is committed to reducing the neonatal mortality rate from 20 per 1,000 live births to the SDG target of 12 per 1,000 live births by 2030.^1^ Addressing gaps in quality of essential and emergency care is a key strategy for achieving this ambitious target.^2–5^ In Mwanza, Tanzania, correct diagnosis estimates at health centers and district hospitals range from 61-87%, while correct treatments are administered only 21-86% of the time.^6, 7^

Provider knowledge is a main driver of care quality and several conventional (in-person) in-service trainings for essential and sick newborn care have been attempted in Tanzania. These include 2^nd^ editions of WHO’s Essential Newborn Care, and the American Academy of Pediatrics’ Helping Babies Breathe.^8, 9^ They range in content scope (care of the newborn in 1^st^ hour vs 1^st^ 28 days of life), levels and mix of cognitive learning targets (i.e. Bloom’s Taxonomy of remembering, understanding, applying, analyzing, evaluating, and creating)^10^ as well as duration of training (2-8 days). Both have demonstrated significant effectiveness when they can be sustained.^11–13^

Unfortunately, conventional in-service education methods are often inadequate in coverage and difficult to sustain.^2, 14, 15^ Conventional education methods do not systematically adapt to individual providers’ knowledge or convenience,^16–19^ have time-limited education and target minimal competency, which limits education effectiveness.^18, 20–24^ Our systematic review highlighted that current educational content and educational design often have limited adaptability to facility needs, which also decreases education effectiveness.^23^ The limited effectiveness and accuracy of current educational methods widens the “know-do” gap, and this gap is greater in rural, under-resourced areas where in-service education is limited.^25^ A key research gap of the World Health Organization is to identify effective provider education that extends across health systems.^26, 27^

Adaptive eLearning, a subdomain of e-learning, pulls from computer science and artificial intelligence principles to create a cognitive model to adapt education to each provider.^28^ Usage data includes metacognition categorized using Howell’s conscious-competence model: 1) conscious competence (correct and confident), 2) unconscious incompetence (confident in knowledge but incorrect), 3) conscious incompetence (not confident in knowledge), and 4) unconscious competence (not confident in knowledge but correct).^29^ Usage data is processed to create a cognitive model for each student and adjusts the sequencing of content and ratio of learning resources based on the formative assessment. Knowledge acquisition during initial learning has been both higher and faster compared to conventional education.28,30

In addition to optimizing initial learning, adaptive eLearning can address forgetfulness through generating refresher learning assignments. Forgetfulness, first described by Ebbinghaus, is an exponential decay of knowledge with knowledge returning to baseline days or weeks after initial learning.^31, 32^ We have seen this decay in our previous work in LMICs with pediatric acute care knowledge and CPR skills.^18, 21^ Forgetfulness can be addressed with refresher assignments, spacing learning over time.^33–35^ Subsequent refresher assignments start at a higher baseline than previous, taking less time to achieve mastery. Over time, yields a more significant percentage of data remembered. In high-income settings, we have demonstrated that refresher assignment completion improves pediatric resuscitation skills and patient outcomes.^36–38^ In Tanzania, the use of refresher assignments over time is a key implementation strategy of Helping Babies Breathe, which has significantly increased adherence to guidelines and reduced neonatal deaths by almost 50%.^13^

Adaptive e-learning has the potential to be rapidly scalable with existing infrastructure: it does not require significant dedicated resources to implement and maintain and may close the training gap that exists for rural, under-resourced areas and allow increased dissemination of up-to date evidence based guidelines and reduce the knowledge deficit, decreasing the need for face-to-face education when instructors are limited.^39–41^ While adaptive e-learning is as effective as conventional education when examining patient outcomes for dyslipidemia screening, monitoring of diabetes, drug dose calculation, and pressure ulcer classification,^42–45^ there is limited evidence examining newborn and pediatric acute care.^46, 47^ Further, the optimal implementation strategy of adaptive e-learning for in-service provider education in low- and middle-income countries (LMICs) is unknown.^45, 48^

Our program, Pediatric Acute Care Education (PACE), is an adaptive e-learning environment co-developed with the Pediatric Association of Tanzania (PAT), Catholic University of Health and Allied Sciences, Stanford University and Area9 Lyceum to improve facility-based adherence to national newborn and pediatric guidelines.(Meaney, digital health 2023) PACE is designed for all cadres (provider types) that may be responsible for caring for newborns and sick children. Initially piloted in 2019, PACE has expanded in size to meet the training needs identified by PAT and PACE providers.

Our initial pilot demonstrated a 30% change in conscious competence during initial learning from 66 (57-75%) to 94% (92-98%).^49^ The average initial learning completion was only 37% and refresher assignment completion was not assessed. There were 3 barriers identified to initial learning completion: 1) use of pre-post assessments for efficacy, 2) lack of in-person technical support, and 3) ineffective email nudging strategy.^49^ Based on these results we revised our implementation strategy in 3 ways: 1) use of change in conscious competence from baseline for efficacy, 2) a full time in-person program manager to provide support PACE providers, and 3) incorporation of an escalating nudge strategy that included emails, WhatsApp messaging and in-person support.

We applied our content development methodology to Tanzania’s essential newborn and sick newborn care guidelines to develop our adaptive Essential Newborn and Sick Care (aESNC) and deployed modules within PACE in May 2022.^50^ aESNC currently contains 2 assignments with total of 9 modules: Preparing for delivery, 1^st^ hour of life, neonatal resuscitation, introduction to sick newborn care, birth asphyxia + pain, convulsions and meningitis, glucose and electrolytes, hemorrhage and jaundice, pneumonia, sepsis, and shock. Learning objectives are restricted to Bloom’s Taxonomy levels 1 and 2 (remembering, understanding), and 100% conscious competence is required to complete the module. Providers completing all PACE modules are awarded continuing professional development credit through PAT toward maintaining professional certification.

The objectives of this study are to assess the implementation success of adaptive Essential and Sick Newborn Care using the revised implementation strategy and describe baseline provider knowledge and metacognition in Mwanza, Tanzania. We evaluated implementation success using the RE-AIM (Reach, Efficacy, Adoption, Implementation and Maintenance) framework adapted by Soicher et al for the implementation of education interventions for higher education and describe provider knowledge and metacognition using Howell’s consciousness competence model.^29, 51, 52^

## METHODS

### Study Design

This was a prospective single-arm, multi-center pilot implementation study conducted in Mwanza, Tanzania from May 2022 to January 2023. This manuscript was formatted in accordance with STROBE guidelines.^53^

### Setting

Study sites included all sites currently participating in PACE that had at least 1 provider in the study cohort. Facility characteristics are listed in Table 1. Due to limited study personnel and this being the initial study, PACE was initially deployed at the zonal hospital in May 2022, and then extended to health centers within Mwanza Region that refer to the zonal hospital in stepwise fashion. Duration of facility participation at time of data extraction is listed in **Table 1**.

**Table 1.** Facility characteristics. *HC: Health Center*

|  | **Zonal**<br>**Hosp** | **HC**<br>**#1** | **HC**<br>**#2** | **HC**<br>**#3** |
| --- | --- | --- | --- | --- |
| **Duration in Study (months)** | 7 | 5 | 4 | 2 |
| **Providers (n)** | 321 | 39 | 66 | 59 |
| **Specialist Care** | + | - | - | - |
| **Pediatrician** | Y | N | N | N |
| **Births/year** | 7000 | 296 | 3678 | 6002 |
| **1m-5y Admissions/year** | 6550 | 0 | 897 | 577 |
| **Services Provided** |  |  |  |  |
| **Outpatient Clinics** | + | + | + | + |
| **Inpatient Wards** | + | - | + | + |
| **Pediatric Ward** | + | - | +/- | + |
| **NICU** | + | - | - | - |
| **Pediatric ICU** | + | - | - | - |
| **Malnutrition unit** | + | - | - | +/- |
| **Cesarean section** | + | - | + | + |
| **Transfusion Services** | + | - | + | + |
| **Pharmacy** | + | + | + | + |
| **Dialysis** | + | - | - | - |

### Participants

The cohort consisted of a convenience sample of consenting healthcare providers who had participated in PACE >30 days. Providers were identified by the facility medical officer in-charge, facility head matron/patron or head of department as well as during sensitization meetings at morning report and recruited to participate by the PACE program manager. Informed electronic consent was obtained through REDCap from all providers who participated in PACE.^54^ Eligible PACE providers are facility-based healthcare providers responsible for providing clinical care to newborns, infants, and/or children. In Tanzania, training duration varies by cadre: medical officers require 5 years of training and 1 year of internship; advanced degree nursing 3 years and 1 year internship; nurses and clinical officers 3 years of training, and assistant medical officers 2 years of training. We excluded providers who declined to consent or who withdrew from study; these numbers are given in the CONSORT diagram, Fig. 1.

### Implementation Strategy

aESNC was implemented through the Pediatric Acute Care Education (PACE) program. PACE is an adaptive e-learning environment with locally derived content, a steering committee, a program manager to provide in-person technical support and series of motivators to increase completion. It is designed to increase provider proficiency in neonatal and pediatric evidence-based guidelines in Tanzania and has been previously described. (Meaney, digital health 2023)

#### Adaptive e-learning with locally derived content

aESNC, consists of 2 assignments: Essential Newborn Care (3 modules, 47 learning objectives) and Sick Newborn Care (6 modules, 91 learning objectives). aESNC content was collaboratively developed with subject matter experts and content designers using Tanzania’s national “Guidelines for Neonatal Care and Establishment of Neonatal Care Unit.^50^ Content creation was supervised by subject matter experts (SME) from the PAT’s Continuing Professional Development Committee (NM, NC, CM), the Tanzanian Society for Pediatric Nursing (ED), and Area9 Senior Learning Architects (BR, MB).

#### PACE Steering committee

The Steering Committee (AH, PAM, AA, HN) provides oversight and coordination of PACE management, research administration, publications and data sharing, and integration of all resources needed for the project. The chair of the steering committee is responsible for communication among committee members, including meeting schedules and agendas, and rotates among the members on a yearly basis. The PACE steering committee consisted of experts in newborn and pediatric care, provider education, and implementation research.

#### The Program Manager

The program manager is a medical (MD) or nursing (RN) officer with experience working in the Tanzanian health system, human subjects research training, effective communication skills and, either formal or informal health education and/or IT skills. The program manager conducts sensitization meetings at facilities and generates contact lists. These lists are reviewed and augmented by the medical officer in charge, head of department and/or chief nursing medical officer as appropriate, and current PACE providers. The program manager meets with consented providers in-person individually to set up PACE on their mobile device, ensure proper functioning and provide initial data bundle.

#### Motivational strategies

- *Nudges*. Weekly, providers who hadn’t completed all PACE initial learning assignments were given reminders or “nudges” to complete their learning. Our escalating nudge strategy was as follows: No activity for 2 days: auto email reminder from Rhapsode; > 5 days: WhatsApp using standardized statements; > 30 days: a virtual or face-to-face meeting with program manager.
- *Internet support*. Providers were reimbursed up to 10gb of data (∼4USD) monthly on their mobile carrier during the study period.
- *Maintaining certification*. Continuing Professional Development credit was awarded via Pediatric Association of Tanzania for completion of all PACE initial learning assignments.
- *Passive feedback to health system leadership*. Program manager would send PACE Facility Progress Report (PDF) to facility stakeholders weekly via email. Stakeholders to this process may include hospital administration, regional and council health management teams, and the Pediatric Association of Tanzania leadership. Reports included aggregate activity, metacognition, and median time to proficiency at a facility level, but not does not report proficiency or metacognition at an individual level.

### Hypothesis

Implementation of aESNC with addition of in-person coordinator and nudging strategy to our implementation strategy would increase reach and improve average completion of both initial learning and refresher assignments compared to our initial PACE pilot.

### Variables

#### Implementation Outcomes

We used the established RE-AIM for the educational intervention implementation framework to define our feasibility outcomes of Reach, Efficacy, and Implementation.^52, 55^ Adoption and Maintenance were not assessed. We assessed Reach using individual participating providers as a proportion of providers identified as eligible.

Efficacy was assessed using change in provider conscious competence (current – baseline). Baseline provider proficiency was determined using providers initial responses to knowledge probes, and current provider proficiency by last response to knowledge probes. Responses were categorized using Howell’s conscious-competence model: 1) conscious competence (correct and confident), 2) unconscious incompetence (confident in knowledge but incorrect), 3)bconscious incompetence (not confident in knowledge), and 4) unconscious competence (not confident in knowledge but correct).^29^

We used 6 metrics to assess implementation: persistent activity, average progress of initial learning completion, average progress of refresher assignment completion, time to enrollment, nudging utilization, and loss to follow-up. Persistent activity was defined as actively using PACE within last 2 weeks of the study (Jan 2023). Average progress of initial learning was percentage of achieving 100% conscious competence of all aESNC learning objectives. Average progress of refresher assignments was percentage of achieving 100% of all content assigned by Rhapsode for refresher learning. Time to enrollment was days from consent to enrollment interview. Nudging utilization included % of providers with at least one nudge, median number of nudges and distribution of nudge types. Loss to follow up was defined as those who were inactive for > 30 days and were not able to be contacted.

#### Predictors, Potential confounders, and Effect modifiers

Several variables were collected to describe the cohort. These included facility, cadre (profession), years of clinical experience, previous newborn training, job satisfaction, motivation, and baseline knowledge and metacognition. Facilities were defined by government designation as a zonal hospital or health center. Cadre categories include specialists, medical officers, assistant medical officers, clinical officers, assistant clinical officers, nursing officers, assistant nurse officers, nurse midwives, medical attendants, laboratory scientist/technologists, and health assistants. The continuous variable “years clinical experience” was collapsed to a categorical variable of <= 1 year, 2-3 years, 4-9 years, and 10+ years based on quartiles rather than visual inspection given small numbers. Previous newborn training was defined as ever having taken either Essentials of Newborn Care, Helping Babies Breathe, both or neither. Job satisfaction and motivation were measured using previous questionnaires validated for healthcare providers in Ghana by Bonenberger *et al*.^56^

### Data sources/measurement

At enrollment, providers completed electronic surveys regarding demographics, previous clinical training, job satisfaction and motivation via REDCap. Provider response data including knowledge competence, metacognition, average progress on initial learning or refresher assignments was collected within the Area9 Rhapsode platform. Data from Rhapsode and REDCap was extracted on January 14^th^, 2023, linked with study ID numbers by name and deidentified prior to analysis.

### Bias

The cohort was identified through sensitization meetings at facilities by the program manager, as well as by facility administrators using staff rosters and study providers and at same points of care over the same time frame for each facility. Outcome data utilized provider usage data from Rhapsode. In addition, enrollment interview and reminders were tracked in the study database. These modules were newly developed, and no study provider had piloted these modules before the study. In addition, baseline competencies were obtained as part of the exposure, allowing to account for an effect of previous training. Our multivariable analyses on factors associated with new provider retention adjusted for plausible prognostic factors. Providers completed all surveys and outcome data electronically using their mobile phones. Potential data entry errors were followed up by the program manager with the provider for clarification where needed and logged within the database. The outcome assessments blinded to the study team and were linked by name and email addresses to the study database and deidentified prior to analysis.

### Study size

As this was a feasibility study, no power calculation was done. Study size was determined by number of providers enrolled during the study period. Based on an estimate of 20% of providers at the zonal hospital and 40% of health center providers caring for newborns and sick children our initial enrollment target was 50 medical officers, 30 clinical officers, and 50 nursing officers over 6 months.

### Statistical methods

Descriptive, univariate, and multivariate analyses were performed. Summary results are presented as means and standard deviations for normally distributed variables and medians with interquartile ranges for variables that were not normally distributed. For our multivariable logistic regression exploring completion of initial learning or persistent activity, we selected provider external characteristics (facility, cadre, experience, previous training) a-priori and did not incorporate response data (job engagement, motivation, or knowledge proficiency data) given the limited size to the cohort. Significance was set at p≤0.05 and models were fit using Python version 3.8.5.

## RESULTS

### Reach

All 4 facilities invited to participate adopted PACE. Of 246 providers, 90% (221) were eligible, with 17 of 25 ineligible due to not completing screening survey. Consent rate of eligible providers was 90% (n=195) with “no time to participate” being the greatest reason for declining participation (**Figure 1**).

### Provider Characteristics

110 (56%) of providers in this cohort were based at the zonal hospital, followed by HC#2 at 42 (22%), HC #1 at 27 (14%) and #3 at 16 (8%). Providers were most commonly medical officers (75, 39%), nursing officers (53, 27%) and clinical officers (21, 11%). Median years of clinical experience was 4 [IQR 1-9]. The reported prevalence of previous newborn training of Essential Newborn Care, Helping Babies Breathe, and both ENC and HBB were 23% was 31%, 41%, and 23%, respectively. Overall job satisfaction was 3.6/5 (SD 0.8) with highest sub scores of morale 4.1/5 (SD 0.8) and supervision 3.8/5 (SD 1.0) and lowest sub scores of renumeration 3.9 (SD 1.2), followed by in-service training 3.5/5 (SD 1.2), management 3.5/5 (SD 1.0) and work environment at 3.5/5 (SD 1.0). Overall motivation scores were 3.7/5 (SD 0.6).

### Efficacy

For providers who completed initial learning (n= 110), average conscious competence increased 42 ± 1 percentage points, while average unconscious incompetence decreased 31 ± 1 percentage points. Of the efficacy, the improvement of conscious competence, 76% was due to reduction in unconscious incompetence. Average conscious incompetence decreased by 10±1 percentage points, while provider unconscious competence was extremely low both before and after PACE. (**Supplementary Data A1-3**).

### Implementation

The average progress of initial learning for all providers was 78% (SD = 31%) (**Supplementary Data C1**). 56% (110) completing all initial learning, 13% (25) incomplete but active within 2 weeks of the end of the study, and 31% (60) becoming inactive during the study period. Providers who completed the initial learning took on average 4.5 [IQR 2.4, 6.1] hours (**Supplementary Data B1-3**). Inactive providers dropped out after on average 1.9 [IQR 0.8, 3.2] hours, and the active providers who are still finishing initial learning spent 4.0 [IQR 2.3, 5.4] hours. Median duration to complete all modules was 61 days [IQR 34,111]. Reduction in probability of provider remaining completing initial learning or remaining active appeared to be evenly distributed over % completion and days of PACE participation (**Supplementary Data C1 and B4**).

Average progress of refresher assignments was 7%, with none completing all refresher assignments, 27% (37) incomplete but active in PACE within last 2 weeks of the study period, and 73% (98) becoming inactive during the study period. Average refresher progress appears to be heavily skewed to lower completion with median completion 2% [IQR:0%, 8%]. (**Supplementary Data C2**). Total hours spent using modules was 0.3±0.7 for active providers and 1.0±1.3 for inactive providers.

Days from consent to enrollment interview was 2 [IQR:0, 4]. 67% (130) of providers needed at least 1 nudge and 30% (59) needed at least one follow-up with the program manager. A total of 1,011 nudges were sent: 311 (31%) email nudges (2-5 days inactivity), 564 (56%) WhatsApp nudges (5-30 days inactivity), and 136 (13%) nudges by the program manager (> 30 days inactivity) (**Supplementary Data D1-2**). The program manager conducted 104 nudges by phone and 30 in-person. Most frequent reasons reported for >30 days inactivity was “no time” (93, 69%), followed by “forgot to use” (68, 50%). Technical barriers (phone not working, can’t access PACE, not sure how to use PACE) represented only 15 (11%) and no provider reported inactivity due to lack of mobile data connectivity. No providers wished to terminate from study when interviewed with program manager at 30 days of inactivity and none were lost to follow-up.

### Baseline knowledge and metacognition of Essential and Sick Newborn Care

Overall median baseline conscious competence for aESNC was 53% [IQR:38-63%], with unconscious incompetence 33%[IQR:25-45%] and 32%[IQR:23-42%] respectively (**Table 3**). aESNC had conscious incompetence of 7% [IQR:2-15%], and unconscious competence 2% [IQR:0-3%].

**Table 2.**
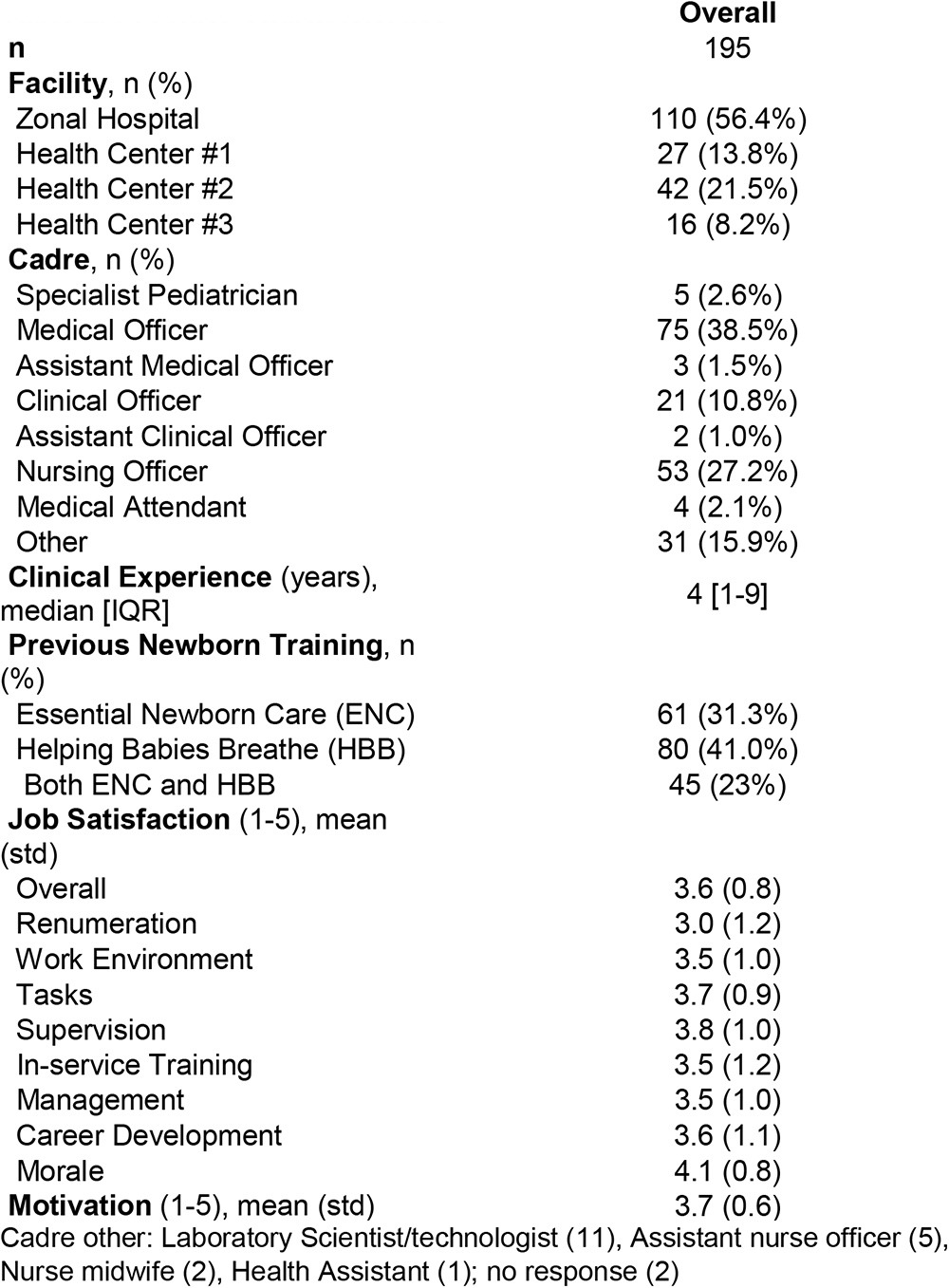
Provider Characteristics.

| **Table 2. Provider Characteristics** | **Overall** |
| --- | --- |
| **n** | 195 |
| **Facility, n (%)** |  |
| Zonal Hospital | 110 (56.4%) |
| Health Center #1 | 27 (13.8%) |
| Health Center #2 | 42 (21.5%) |
| Health Center #3 | 16 (8.2%) |
| **Cadre, n (%)** |  |
| Specialist Pediatrician | 5 (2.6%) |
| Medical Officer | 75 (38.5%) |
| Assistant Medical Officer | 3 (1.5%) |
| Clinical Officer | 21 (10.8%) |
| Assistant Clinical Officer | 2 (1.0%) |
| Nursing Officer | 53 (27.2%) |
| Medical Attendant | 4 (2.1%) |
| Other | 31 (15.9%) |
| **Clinical Experience** (years),<br>median [IQR] | 4 [1-9] |
| **Previous Newborn Training, n (%)** |  |
| Essential Newborn Care (ENC) | 61 (31.3%) |
| Helping Babies Breathe (HBB) | 80 (41.0%) |
| Both ENC and HBB | 45 (23%) |
| **Job Satisfaction** (1-5), mean<br>(std) |  |
| Overall | 3.6 (0.8) |
| Remuneration | 3.0 (1.2) |
| Work Environment | 3.5 (1.0) |
| Tasks | 3.7 (0.9) |
| Supervision | 3.8 (1.0) |
| In-service Training | 3.5 (1.2) |
| Management | 3.5 (1.0) |
| Career Development | 3.6 (1.1) |
| Morale | 4.1 (0.8) |
| **Motivation** (1-5), mean (std) | 3.7 (0.6) |
| Cadre other: Laboratory Scientist/technologist (11), Assistant nurse officer (5),<br>Nurse midwife (2), Health Assistant (1); no response (2) |  |

**Table 3.**
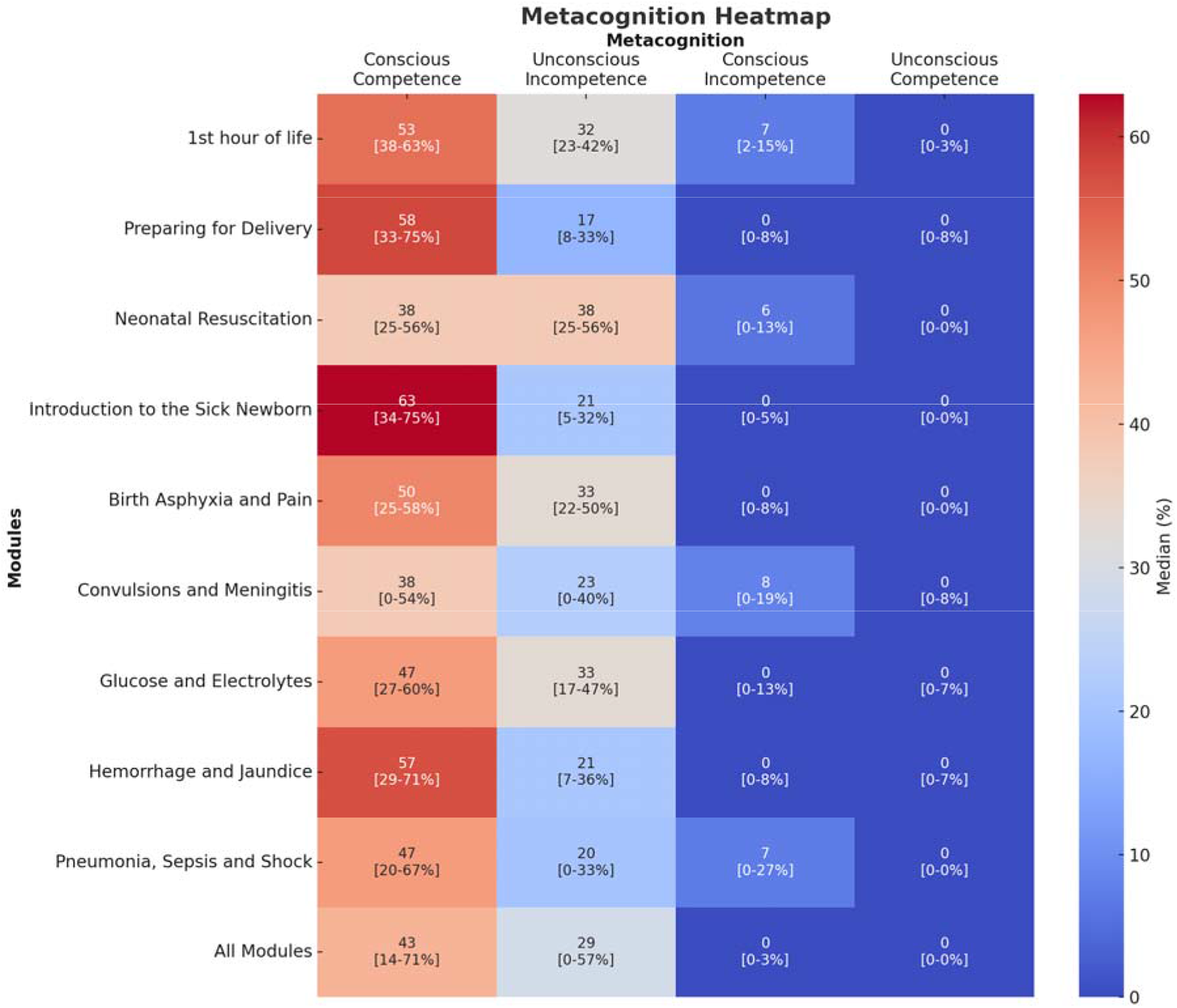
Baseline Metacognition Heatmap.

For individual aESNC modules, conscious competence was highest in Neonatal Resuscitation (63% [IQR:34-75%]), 1^st^ Hour of Life (58% [IQR:33-75%]), and Glucose and Electrolytes (57% [IQR:29-71%]), and lowest in Birth Asphyxia and Pain (38% [IQR:0-54%]), Preparing for Delivery (38% [IQR:25-56%]), and Pneumonia, Sepsis and Shock (43% [IQR:14-71%]). Unconscious incompetence was highest in Preparing for Delivery (38% [IQR:25-56%]), Introduction to the Sick Newborn (33% [IQR:22-50%]), and Convulsions and Meningitis (33% [IQR:17-47%]), and lowest in 1^st^ Hour of Life (17% [IQR:8-33%]), Hemorrhage and Jaundice (20% [IQR:0-33%]), Glucose and Electrolytes (21% [IQR:7-36%], and Neonatal Resuscitation (21% [IQR:5-32%]). Conscious incompetence was highest in Birth Asphyxia and Pain (8% [IQR:0-19%]), Hemorrhage and Jaundice (7% [IQR:0-27%]) and Preparing for Delivery (6% [IQR:0-13%), and zero in all others ([IQR:0-13%]). Unconscious competence was zero in all modules ([IQR:0-8%]).

### Factors associated with initial learning completion or persistent activity

On univariate analysis, being a nursing officer, 2-3 years of experience, and having higher baseline conscious competence was associated with initial learning completion or persistent activity. On multivariate modeling, nursing officers, clinical officers and “other” personnel were significantly associated with better initial learning completion or persistent activity when compared to medical officers adjusting for facility, clinical experience, and previous newborn training (Table 4). Facility, previous training with either Essential Newborn Care or Helping Babies Breathe were not associated with initial learning completion or persistent activity.

**Table 4.** Factors associated with initial learning completion or persistent activity.

|  | Univariate Odds Ratio<br>[95% CI) | Adjusted Odds Ratio<br>[95% CI) |
| --- | --- | --- |
| **Facility** |  |  |
| Zonal Hospital | comparison | comparison |
| Health Center #1 | 1.6 [0.6, 4.2] | 0.5 [0.1, 2.4] |
| Health Center #2 | 0.8 [0.4, 1.7] | 0.3 [0.1, 1.3] |
| Health Center #3 | 1.0 [0.3, 3.0] | 0.5 [0.1, 2.5] |
| **Cadre** |  |  |
| Specialist Pediatrician | insuff. data | insuff. data |
| Medical Officer | comparison | comparison |
| Assistant Medical Officer | insuff. data | insuff. data |
| Clinical Officer | 1.0 [0.4, 2.6] | 2.4 [0.5, 11.2] |
| Assistant Clinical Officer | insuff. data | insuff. data |
| **Nursing Officer** | **3.4 [1.4, 8.1]** | **5.6 [1.8, 18.1]** |
| Medical Attendant | insuff. data | insuff. data |
| Other | 1.1 [0.5, 2.6] | 2.4 [0.6, 9.6] |
| **Clinical Experience** |  |  |
| ≤ 1 year | comparison | comparison |
| **2 - 3 years** | **0.3 [0.1, 0.8]** | 1.3 [0.4, 4.7] |
| 4 - 9 years | 0.7 [0.3, 1.8] | 0.3 [0.1, 1.1] |
| ≥ 10 years | 1.1 [0.4, 2.8] | 0.8 [0.3, 2.1] |
| **Previous Newborn Training** |  |  |
| Essential Newborn Care (ENC) | 1.0 [0.5, 1.9] | 1.4 [0.6, 3.4] |
| Helping Babies Breathe (HBB) | 1.0 [0.5, 1.8] | 0.6 [0.2, 1.5] |
| **Job Satisfaction (1-5)** |  |  |
| Overall | 1.0 [0.7, 1.4] | - |
| Remuneration | 1.1 [0.8, 1.4] | - |
| Work Environment | 1.0 [0.7, 1.3] | - |
| Tasks | 0.7 [0.5, 1.0] | - |
| Supervision | 1.0 [0.8, 1.4] | - |
| In-service Training | 1.1 [0.9, 1.5] | - |
| Management | 1.0 [0.8, 1.4] | - |
| Career Development | 1.0 [0.7, 1.3] | - |
| Morale | 1.1 [0.8, 1.7] | - |
| **Motivation (1-5)** | 1.2 [0.7, 2.1] | - |
| **Baseline Scores (0-1)** |  |  |
| **Conscious Competence** | **31.6 [5.8, 183.5]** | - |
| Unconscious Competence | 1.4 [0.1, 14.6] | - |
| Conscious Incompetence | insuff. data | - |
| Unconscious incompetence | 0.9 [0.1, 5.8] | - |

## Discussion

This study demonstrated that adaptive eLearning can increase reach to providers not reached by conventional training, and the use of in-person support and an escalating nudging strategy increased initial learning completion. Further, refresher assignment completion was low, and our implementation strategy needs to be revised to solidify knowledge gained through initial learning. Finally, provider awareness of knowledge gaps was low at baseline and lower baseline knowledge is associated with decreased completion or learning activity.

aESNC reached 85% of providers in this study. The critical mass needed for collective behavior change for providers in facilities is unknown, but using estimates from current social behavioral science suggest that 10-40% of providers are needed to be engaged to drive behavior change.^57–59^ As knowledge is only one component that drives behavior change and we know not all providers would complete all modules, we will examine thresholds for reach required to achieve sustained changes in quality of care.

aESNC was able to reach providers that conventional newborn training had not reached. We found that 30% of providers had received Essential Newborn Care training, 40% had received Helping Babies Breathe, but only 23% had received both. The impact of this limited reach may be underestimated if this reported training was years ago. A reason for conventional education’s limited reach may be due to high turnover of front-line providers at academic centers, which may be able sustain training but not sufficient to address its training gaps. Furthermore, community health centers may have limited support to hold local training or send staff for training to meet this need. This conclusion from this initial study should be tempered as our observed reach, as well as our enrollment rates, were higher than anticipated, and don’t correlate with staffing estimates reported from health system leadership (Table 1). This may be due to a combination of incomplete lists by facility leadership and early adopter phenomenon. Future studies will bear this out.

Our initial efficacy of 41% was consistent with our previous conventional studies that demonstrated knowledge efficacy rates of 10-25% after pediatric acute care training using current training methods^18–,21^, as well as other newborn and pediatric conventional in-service education studies.^5, 22^ It is important to note that our use of formative assessments at baseline and during the educational session, are relatively new measures of educational efficacy. In our pilot, the difference between latest conscious competence and baseline conscious competence was roughly 10 points higher than between pre-post testing.^49^

The average progress of providers for initial learning was 78%, and 56% of providers completed all initial learning assignments, higher than we observed in our initial pilot of PACE. This may be due to the use of in-person support and an escalating nudging strategy increased initial learning completion. The completion rate in this study is comparable to previous studies in high income countries: much higher than massive open online courses progress rates of 7-15%,^60, 61^ and comparable to longitudinal e-learning studies of healthcare providers (44-50%).^62, 63^ A few shorter courses (< 3 months) with more in-person support report completion rates as high as 75-80% without incentives,^64, 65^ and as high at 70-96% completion when incentive of $200-$750 was provided.^66, 67^

In this study, we observed more time is needed for providers to complete initial learning than anticipated. We had estimated providers needing 5 hours over 1 month to complete initial learning. While median time to complete initial learning was shorter than what we targeted (4.5 hours), the median days to completion of initial learning was 61 days, twice what we had estimated. Interestingly, the retention curve demonstrated no significant drop-off that had been seen previously. Inadequate time for education, not technical issues or lack of internet access was the main barrier.

Other reasons, besides the in-person support and escalating nudging strategy, may have contributed to these findings. We did not use a designated order determined by subject matter experts as we did in our initial pilot, as we observed a significant drop off with disability and exposure assessment. This choice of module completion may have allowed better alignment of provider learning interest and increased engagement during initial usage, and familiarity with adaptive learning may have facilitated usage of completion of additional content. Additionally, there may have been increased peer support for adaptive learning due to a higher percentage of providers participating in PACE at each facility. Whether this improved average completion is due to our implementation strategy components (escalating nudging strategy, program manager, reports to facility), changes to PACE itself (self-selection of topics), or environmental (unmeasured peer support) is unclear.

Our implementation strategy needs further revision to increase refresher assignment completion. Less than a third of participants participated in refresher assignments, and the average progress through refresher assignments was <10%. Improving established provider retention and completion of refresher assignments is key to optimizing education effectiveness. In-service provider education is designed to build upon foundational knowledge gained in pre-professional training, as well as solidify it through relevance to a provider’s clinical experience. Unfortunately, many continuing professional education programs are short courses without longitudinal follow-up. The use of in-person skills training, tailored to provider needs using peers as “clinical champions”, may enhance refresher assignment completion. These clinical champions may add a point of engagement and facilitate normalization of adaptive eLearning use to facilitate education over time. Additionally, as our nudging strategy was triggered only to complete all initial learning assignments, providers may believe they have no more learning to do. Further refinements with our nudging strategy and sensitization meetings with facility providers and leadership to emphasize importance of refresher training may also help normalize adaptive e-learning for providers.

Adverse events occur in 18% of inpatient admission in Africa and continuous learning is needed to improve patient safety.^68, 69^ Although baseline knowledge of guidelines was consistent with previous studies, we identified that providers are often unaware of knowledge gaps. In addition to demonstrating that providers know about half of newborn knowledge at baseline, adaptive e-learning allows us to see providers’ awareness of their knowledge and misconceptions. Unconscious incompetence is a latent threat to health systems, and as they are not realized, may mask workforce knowledge gaps. Unconscious incompetence is a particular danger clinically, as providers are confident but incorrect in what care they believe should be delivered.

While aESNC increased provider knowledge, our exploratory analysis identified that baseline lower knowledge may be associated with inactivity or non-completion of aESNC. While one of the strengths of adaptive learning is using formative assessments to facilitate learning, providers whose baseline knowledge is too low may suffer cognitive overload. Our level of unconscious incompetence surpassed Area9’s best practice guidelines, which suggest keeping it between 20 and 30%. It was also higher than what we observed in our initial pilot. Development of a strategy of focused support to engage with those at risk may be needed.

### Limitations

Several limitations are important to note. First, job satisfaction and motivation has not been validated for turnover intention in Tanzania. Also, our study had limited ability to monitor implementation metrics. Despite having solid relationships and regular meetings, we faced significant challenges in obtaining complete lists of trained providers. This issue could have non-randomly limited our ability to reach less motivated or disengaged providers. Finally, our univariate and multivariable analyses were limited by numbers and may have failed to detect important provider characteristics that impact provider retention.

### Future directions

All the metrics generated need to be validated against clinically meaningful outcomes across various contexts. Adaptive e-learning may allow us to better understand the how, when why and where providers learn, as well as how that knowledge is translated into behavior change and improved patient outcomes. There is also a need for user input in defining what is essential to optimize effectiveness when fed back to providers and healthcare managers for quality improvement. A better understanding of how providers learn will guide the development of education implementation strategies that are not only effective, but scalable and sustainable. Future studies should improve the identification of all eligible providers, monitor the use of data bundles, fidelity of escalating nudge strategy to individual providers, aggregate reports to facility leadership, and distribution of CPD points.

The immediate next steps are to conduct qualitative research to identify barriers and facilitators to adaptive eLearning and refine our implementation strategy to include tailored skills training and local capacity to develop new content continuously. In addition, we will refine our clinical auditing methodology to validate learning metrics on clinically meaningful outcomes.

## Conclusion

aESNC reached many providers without previous conventional in-service training in both sick and essential newborn care. Use of in-person support and motivators increased initial learning completion, but refresher assignment completion was low. Providers were often unaware of knowledge gaps, and lower baseline knowledge may be associated with non-completion of initial learning. Further study examining tailored skills trainings, clinical champions, continuous content development, as well as qualitative studies to identify barriers and facilitators to normalizing adaptive e-learning in provider behavior is needed.

## Supporting information

STROBE Checklist

## Data Availability

All data produced in the present study are available upon reasonable request to the authors.

## Acknowledgements

The authors would like to thank the Pediatric Association of Tanzania; the Tanzanian Ministry Of Health, Regional and Council Health Management Teams for participating in stakeholder meetings, Géraldine Jossellin-Duval for her leadership and mentorship of the PACE Learning Engineering Team; Denis Albert and Agnes Hassan for their organizing of participants and site work in Mwanza; Michael Alfonzo, Jose “Jojo” Ferrer, Segolame Setlhare, CLN mentors and facilitators for their valuable feedback on the initial versions of aESNC and its refinements.

**SAGER guidelines for sex and gender reporting were followed.**

**Consensus statement on measures to promote equitable authorship.**

## Contributors

**Substantial contributions by author:**

| Authors | Study<br>Conception/design | Data |  |  |
| --- | --- | --- | --- | --- |
|  |  | Acquisition | Analysis | Interpretation |
| Peter A Meaney | Yes | Yes | Yes | Yes |
| Adolfine Hokororo | Yes | Yes | Yes | Yes |
| Hanston Ndosi | Yes | Yes | Yes | Yes |
| Alex Dahlen | Yes | No | Yes | Yes |
| Theopista Jacob | Yes | No | No | Yes |
| Joseph R Mwanga | No | No | No | Yes |
| Florence S Mukalaba | No | No | No | Yes |
| Christine Joyce | No | No | No | Yes |
| Rishi Mediratta | No | No | No | Yes |
| Boris Rozenfeld | Yes | No | No | Yes |
| Marc Berg | Yes | No | No | Yes |
| Zack Smith | Yes | No | No | Yes |
| Neema Chami | No | No | No | Yes |
| Namala P Mkopi | No | No | No | Yes |
| Castory Mwanga | No | No | No | Yes |
| Enock Diocles | No | No | No | Yes |
| Ambrose Agweyu | Yes | No | Yes | Yes |

All Authors drafted the work or revised it critically for important intellectual content; AND approved the final version to be published; AND agree to be accountable for all aspects of the work in ensuring that questions related to the accuracy or integrity of any part of the work are appropriately investigated and resolved.

## REDCap Database

Study data were collected and managed using REDCap electronic data capture tools hosted at Stanford University.^70, 71^ REDCap (Research Electronic Data Capture) is a secure, web-based software platform designed to support data capture for research studies, providing 1) an intuitive interface for validated data capture; 2) audit trails for tracking data manipulation and export procedures; 3) automated export procedures for seamless data downloads to common statistical packages; and 4) procedures for data integration and interoperability with external sources. The Stanford REDCap platform (http://redcap.stanford.edu) is developed and operated by Stanford Medicine Research IT team. The REDCap platform services at Stanford are subsidized by a) the Stanford School of Medicine Research Office, and b) the National Center for Research Resources and the National Center for Advancing Translational Sciences, National Institutes of Health, through grant UL1 TR001085. Area9 Rhapsode™ meets the requirements for full GDPR compliance including encryption, data security, and ‘forget me’.

## IRB language

The Institutional Review Board of the Tanzania National Institute of Medical Research (NIMR/HO/R.8a/Vol.IX/3990), Stanford University (60379), the ethics committee of the Catholic University of Health and Allied Science (no ID number given), and the Mwanza Regional Medical Officer (Ref. No. AG.52/290/01A/115) approved the study protocol including consent procedures. Data collection procedures were completed in compliance with the guidelines of the Health Insurance Portability and Accountability Act (HIPAA) to ensure subject confidentiality. Informed electronic consent was obtained through REDCap from all providers who participated in PACE.^54^ All providers who completed consent were included. All surveys and questionnaires were entered directly by providers into REDCap. This study is reported according to the Consolidated Standards of Reporting Trials (CONSORT) 2010 extension to randomized pilot and feasibility trials.

## Patient and public involvement

This research was done without patient involvement. Patients were not invited to comment on the study design and were not consulted to develop patient-relevant outcomes or interpret the results. Patients were not invited to contribute to the writing or editing of this document for readability or accuracy.

