## Supplemental Tables for "Feasibility of an Adaptive E-Learning Environment to Improve Provider Proficiency in Essential and Sick Newborn Care in Mwanza, Tanzania"

Supplementary Materials:

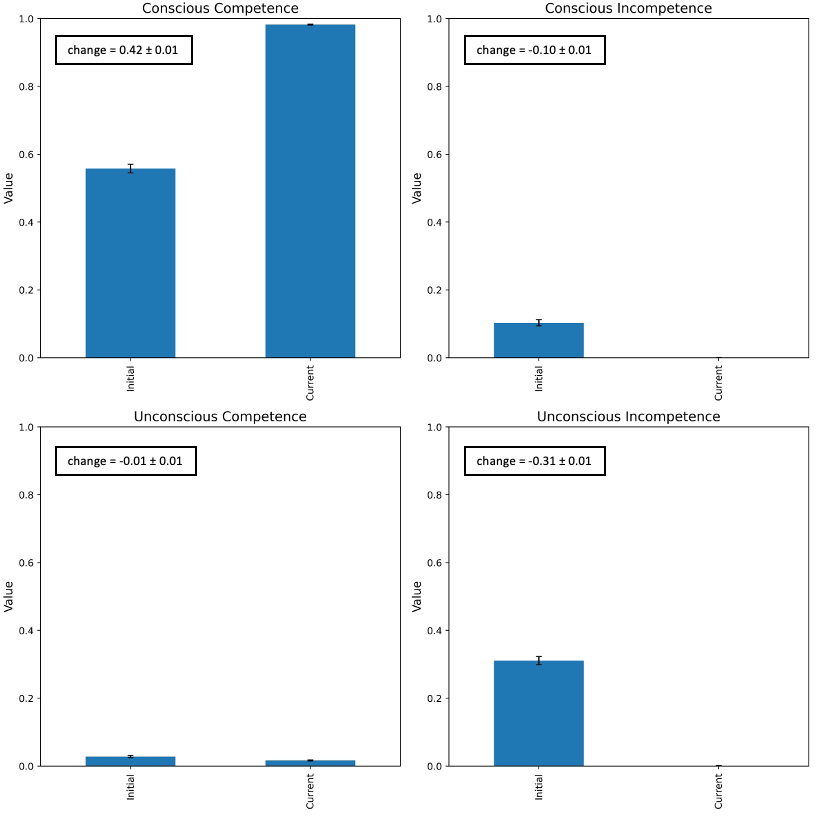

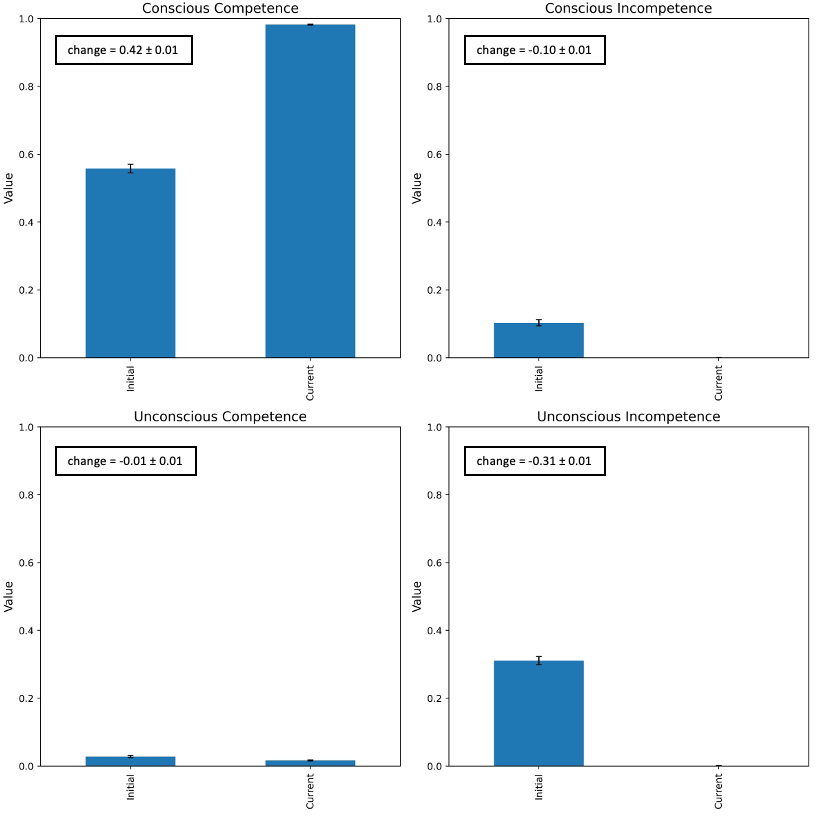

**Figure A. aESNC Efficacy.**

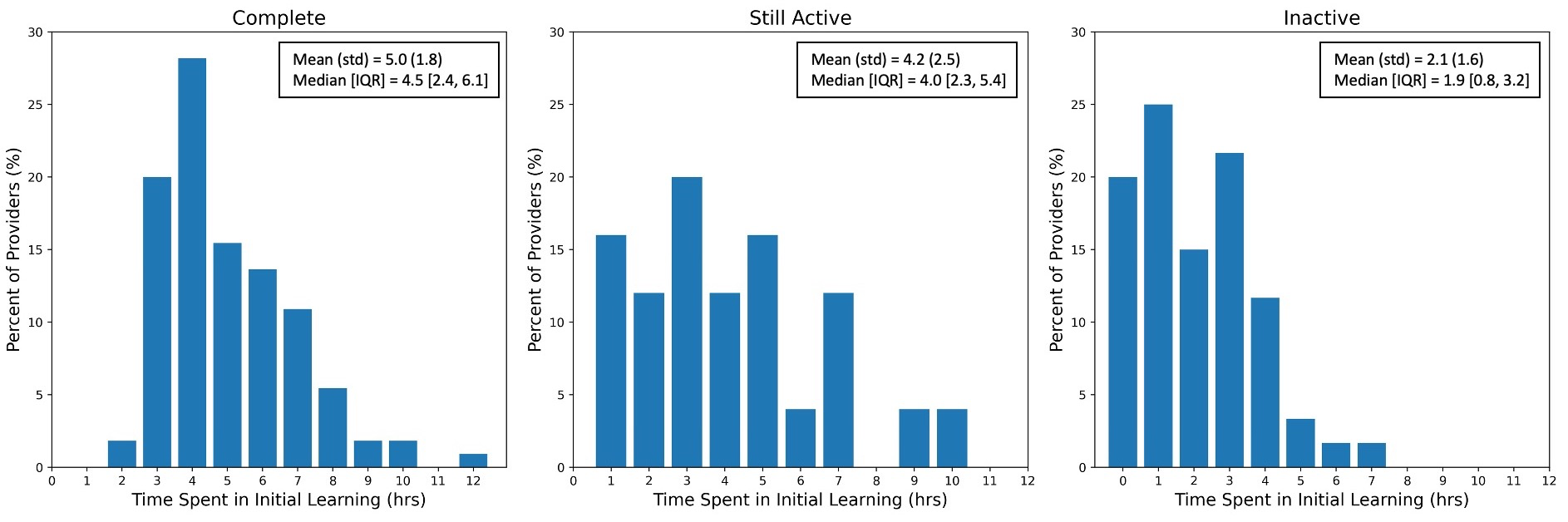

**Figure B1	.	Figure B2.	Figure B3.**

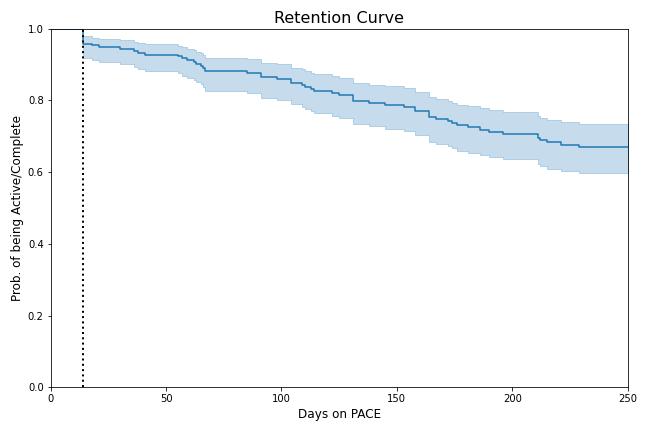

**Figure B4.**

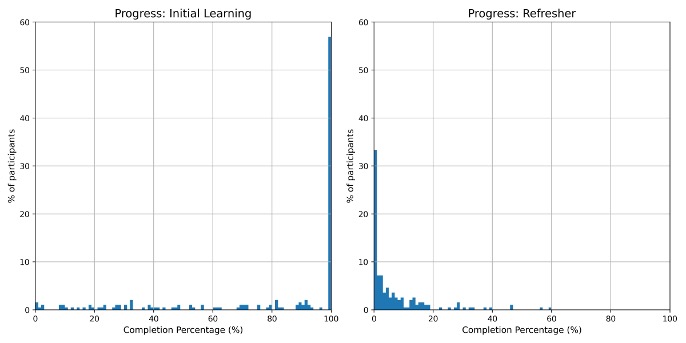

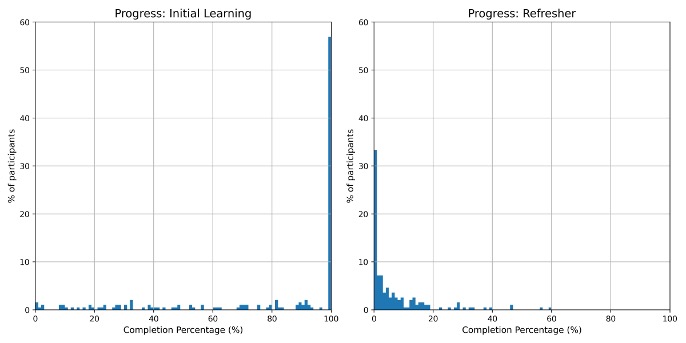

**Figure C1, C2. Progress Distribution**

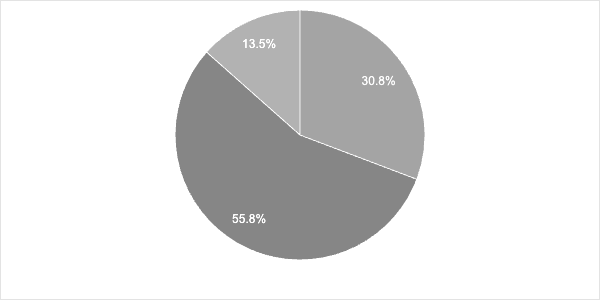

**Figure D1: Nudge types**

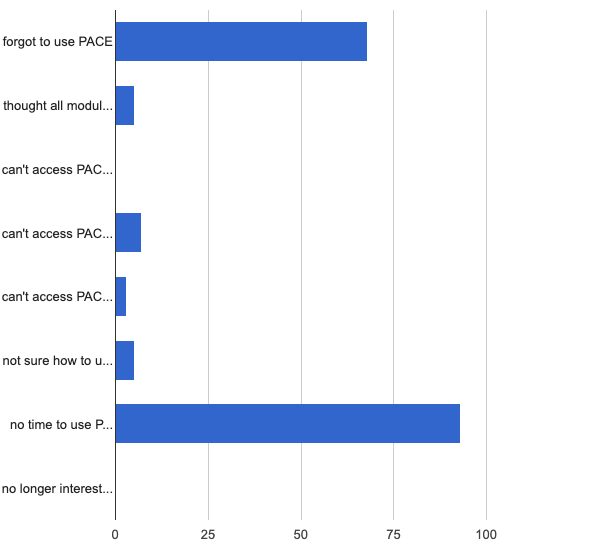

**Figure D2: Reasons for > 30days inactivity**

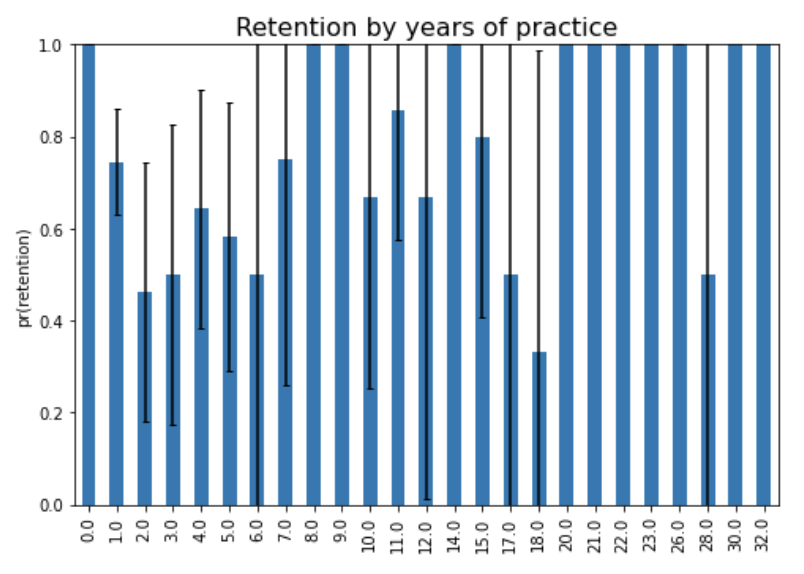

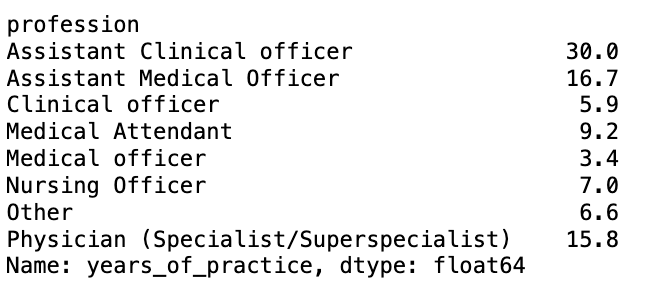

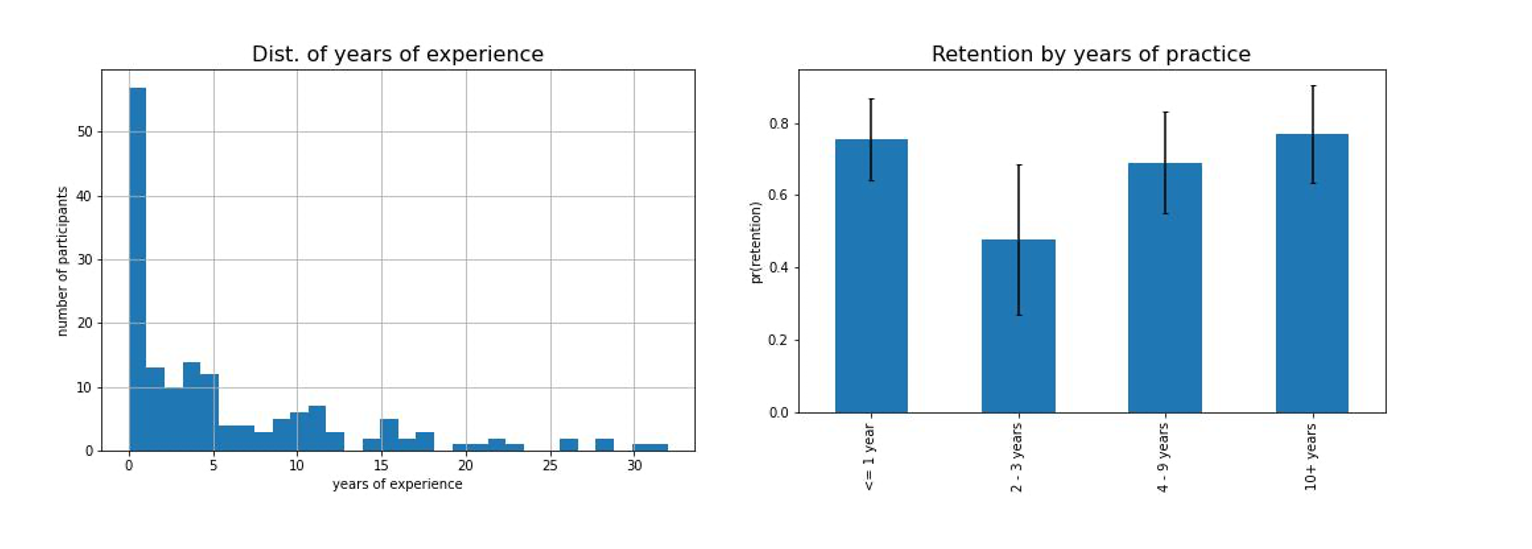

**Figure. Years of experience**

*Providers with 2-3 years of experience have the lowest retention rates.*
